# PHeP: TrustAlert Open-Source Platform for Enhancing Predictive Healthcare with Deep Learning

**DOI:** 10.1101/2024.08.22.24311205

**Authors:** Sandro Gepiro Contaldo, Emanuele Pietropaolo, Lorenzo Bosio, Simone Pernice, Irene Terrone, Daniele Baccega, Yuting Wang, Rahul Kumar Sahoo, Giuseppe Rizzo, Alessia Visconti, Paola Berchialla, Marco Beccuti

**Affiliations:** Department of Computer Science, University of Turin, Turin, Italy; Laboratorio InfoLife, Consorzio Interuniversitario Nazionale per l’Informatica (CINI), Italy; Centre for Biostatistics, Epidemiology and Public Health, Department of Clinical and Biological Sciences, University of Turin, Turin, Italy; Polytechnic of Turin, Turin, Italy; LINKS Foundation, Turin, Italy

**Keywords:** predictive healthcare, deep learning, high performance computing, natural language processing, medical informatics

## Abstract

Predictive healthcare, a field revolutionized by the availability of large medical datasets and computational advancements, plays a pivotal role in enhancing patient care and healthcare system performance. Despite its significance, the training and deployment of predictive healthcare models present substantial challenges. To address these, we introduce the Predictive Healthcare Platform (PHeP), an open-source platform that simplifies the use of pretrained models. PHeP, developed under the TrustAlert project, offers an intuitive web-based graphical interface, making advanced predictive healthcare accessible to users without extensive computational skills or expensive computational resources. This paper presents the design and functionality of PHeP, demonstrating its effectiveness in predictive healthcare tasks, such as predicting re-hospitalization at three months using pre-trained BERT models.

## 1 Introduction

Predictive healthcare, *i.e*., the use of historical and/or real-time data to anticipate patients’ medical needs, is crucial to improve both patients’ quality of life and healthcare systems’ performances. It has become possible thanks to the availability of large datasets of medical records, such as those collected in electronic health records (EHRs) and healthcare administrative databases (HADs), and to the recent development in deep learning (DL) approaches. EHRs and HADs track patients’ medical history by collecting longitudinal information on medical diagnoses and procedures, as well as drug prescriptions. DL approaches are naturally able to deal with these very large amounts of longitudinal data, also when presenting with irregular time intervals between events [1]. These types of techniques have been successfully used to predict the risk of hospitalization, the onset of new diseases, and the worsening of existing ones [1, 2, 3]. Performances of DL approaches can be further improved by combining them with natural language processing (NLP) techniques, which add contextual information by generating a compact representation of medical data (embedding). This line of research has been spurred by the development of the Bidirectional Encoder Representations from Transformers (BERT) [4]. BERT allows pre-training deep bidirectional representations from unlabelled text, which can be then fine-tuned to solve a wide range of tasks, including predictive healthcare tasks [5, 6, 7], without substantial architecture modifications.

Training BERT and its derived models, however, requires a very large amount of data, nontrivial expertise in data munging and DL technologies, state-of-the-art and costly hardware, and long processing time. For instance, training the large model (BERTLARGE) took 4 days on 16 Cloud TPUs (64 TPU chips total) [4] and training of Med-BERT [7] took a week on a single Nvidia Tesla V100 GPU of 32GB graphics memory capacity.

To address this and enhance the adoption of DL approaches in healthcare, we have developed a new open-source Predictive Healthcare Platform (PHeP). PHeP provides access to pre-trained models through an intuitive web-based graphical interface, aiming to simplify their usage for users without advanced computational skills. Here, we describe PHeP and demonstrate its effectiveness creating an online service to predict re-hospitalization at three months using two pre-trained BERT models.

## 2 Methods

This section outlines the main subcomponents of PHeP, details the hardware and software stack supporting it, and describes the pre-trained models used in our case study.

### 2.1 PHeP in a nutshell

PHeP sports a modular architecture with four main subcomponents: Collaborative Working Environment, Data Repository, Analysis Environment, and Prediction Environment (Figure 1A).

**Figure 1:**
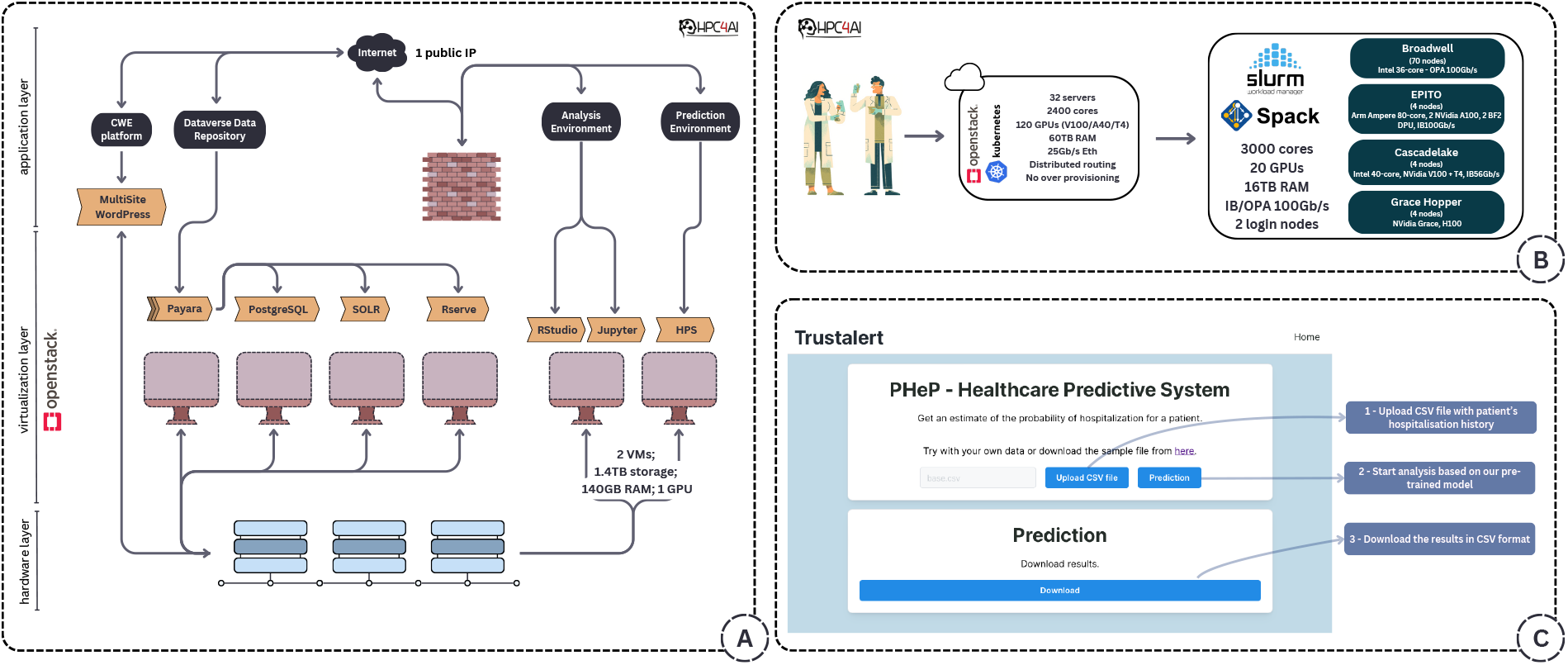
(A) Overview of the hardware and software stack supporting PHeP. Black boxes represent PHeP sub-components, orange boxes represent micro-services for each sub-component, and arrows illustrate their interdependencies. (B) The open-access High-Performance Computing for Artificial Intelligence (HPC4AI) architecture. (C) The PHeP user interface for our case study

The *Collaborative Working Environment* (CWE, (https://www.trustalert.it) acts as the front-end platform, and serves as a centralized hub, allowing users to interact with various PHeP services, access resources, and engage with TrustAlert Project-related information [8]. Users can seamlessly navigate functionalities, collaborate with team members, access project documentation, monitor progress, and stay updated on developments. It is implemented through a WordPress multisite instance which allows for the creation of multiple sites within a single WordPress installation. Notably, the WordPress multisite instance was intentionally deployed in a separate virtual network from the other services to mitigate the potential impact of a security breach within WordPress on critical services. All connections between the CWE and the other primary services undergo a firewall service, thus effectively filtering malicious traffic.

The *Data Repository* is a centralized location providing a secure and structured environment, for storing, organizing, and managing data produced in the project, according to FAIR principles, including all pre-trained models developed for predictive purposes. It is implemented through Harvard Dataverse [9], a leading open-source data repository platform designed to facilitate the sharing, preservation, and discovery of research data. The repository offers persistent identifiers and version control features, ensuring the integrity and traceability of data over time. This facilitates the reuse and validation of data by other researchers, and supports reproducibility in scientific research. The Data Repository can interact with the other subcomponents through specifically developed REST APIs.

The *Analysis Environment* provides TrustAlert researchers with access to two powerful and versatile data analysis environments, *i.e*., R and Python. The R environment is based on RStudio Server, which offers a robust and user-friendly interface for R programming and statistical analysis. The Python environment is hosted on JupyterHub, providing an interactive platform for Python coding, data visualization, and machine learning tasks. Thus, the *Analysis Environment* offers the necessary tools and resources for tasks such as preprocessing data, conducting exploratory data analysis, and building predictive models.

The *Prediction Environment* is the cornerstone of PHeP, embodying its cutting-edge Healthcare Predictive System (HPS). Through a seamlessly integrated web interface and an encrypted communication channel, this environment grants users intuitive access to a suite of advanced and secure predictive analytics tools. It can be easily instantiated with various pre-trained models, and customized to address different predictive healthcare tasks, such as the risk of short-term hospitalization (already implemented), the onset of new diseases, and the risk of non-urgent A&E accesses. Within the *Prediction Environment*, we developed a specialized web interface tailored for executing predictions via Python scripts. Harnessing the power of Next.js, a robust React framework renowned for its ability to construct high-quality web applications with server-side rendering and static generation, we streamlined the development process and expedited deployment, resulting in an intuitive and responsive platform. By using Next.js, we optimized code management and bolstered code reusability. The framework lazy loading strategy and automatic code-splitting efficiently render pages, loading only necessary content to enhance performance by fetching precisely what is required.

### 2.2 Hardware and software stack supporting PHeP

The hardware infrastructure hosting PHeP relies on the open-access High-Performance Computing for Artificial Intelligence (HPC4AI) data center (https://hpc4ai.unito.it), situated at the University of Turin. HPC4AI introduces an innovative HPC-cloud convergence architecture (Figure 1B), reshaping the traditional utilization of cloud and HPC systems to bolster AI applications. In this model, the cloud provides a modern interface for HPC, and HPC acts as an accelerator for the cloud. The HPC4AI cloud system is implemented using the open-source OpenStack cloud technology [10], and leverages different resources, including over 2400 physical cores, 60 TB RAM, 120 GPUs (NVidia T4/V100/A40), 25 Gb/s networking, and 4 storage classes with distinct characteristics. The use of OpenStack facilitates the implementation of a robust Deployment-as-a-Service (DaaS) feature, allowing the modeling of user-defined platforms (IaaS and PaaS) in a portable manner. Moreover, it automates their deployment on a virtualized infrastructure. The HPC subsystem is managed through SLURM software [11] and comprises 72 nodes with Intel architecture (68 nodes with 32 cores, 128GB RAM, OPA 100Gb/s and 4 node with 40 cores, 1 TB RAM, T4+V100 GPUs, IB 56Gb/s) and 4 nodes with Arm architecture (Ampere Altra 80 cores, 512GB RAM, 2xA100 GPUs, 2xBF2 DPUs, IB 100Gb/s). Additionally, there are 2 HPC storage systems utilizing BeeGFS [12] and LUSTRE [13], both in an all-flash configuration.

Figure 1A show an overview of the hardware and software stack supporting PHeP. Above the hardware layer, hosted within the HPC4AI data centre, lies the virtualization layer, powered by the OpenStack project, and which encompasses two Virtual Machines, collectively offering 20 Virtual Cores, 140GB RAM, and 1.4TB of storage. At the top is the application layer, which includes three of the integrated subcomponents (black boxes) described in the previous subsection, along with its corresponding microservices (orange boxes). The connections between these components are visually depicted by arrows, illustrating their interdependencies. This layered architecture ensures robust and scalable performance. The hardware provides a solid foundation, while virtualization efficiently facilitates resource management.

### 2.3 Proof-of-concept: BERT for predicting short-term re-hospitalisation

The dataset was created from an anonymized real data set, in which the information were reshuffled and recombined conditioning on both age class and sex. The real data used in this study were collected by one of the local health services in the Piedmont region of Italy (ASL-CN2). The ASL-CN2 Ethics Committee and the Data Protection Officer authorized this study (Presa d’atto n. 4-2023). The simulated dataset can be downloaded from http://trustalert.hpc4ai.unito.it:3000/.

To exploit the multi-head self-attention and positional encoding of transformer-based architectures of BERT, we chronologically sorted the hospitalisation events (sentences), each including a variable number of importance-sorted diagnoses (words) to define a patient’s medical history (document).

We divided our dataset in training, including 80% of the data, and testing set. We then pretrained BERT both on the Masked Language Modelling (MLM) and Next Sentence Prediction (NSP) tasks originally proposed [14] (*BertForPreTraining* implementation; 8 epoch). The MLM task randomly masks a portion of the input sentences, and then learns to predict it based on the surrounding context, effectively capturing the relationships between same-hospitalisation diagnoses. The NSP tasks extract random pairs of sentences and classify whether these are consecutive (*i.e*., follows each other chronologically), thus learning temporal connections between hospitalisation events and their associated diagnoses. We generated two pre-trained models by training a raw version of a model architecture only on our HAD data (*baseline*) and by further pre-training the *bert-base-uncased* version trained on the English language database again using our HAD data (*English-HAD*)

BERT’s ability to create meaningful embeddings is usually evaluated with downstream tasks such as question answering, entity recognition, and text summarisation. Here, we predicted whether a patient will be re-hospitalised within three months given their medical history (*BertForSequenceClassification* implementation; 1 epoch). In details, for each patient with at least two hospitalisations, we removed the last hospitalisation from the dataset, using it to label the patient’s medical history as ‘1’ if it happens within three months from the previous one, or ‘0’ otherwise. To increase the number of available sentences, out of every patient’s clinical history, we generated *n*_*h*_ − 1 labelled sequences (where *n*_*h*_ is the number of hospitalisation events).

Due to the proof-of-concept nature of this study, we did not performed any hyper-parameters tuning, and the values selected from a literature review for both pre-training and fine-tuning are shown on the PHeP website.

## 3 Results

To validate the capabilities of the PHeP system, we conducted a comprehensive series of tests and experiments. The initial experiment aimed to demonstrate how PHeP can efficiently exploit the hardware resources available through the HPC-cloud convergence infrastructure, highlighting its ability to scale. In detail, we utilized simulated data of varying sample sizes (N=12K, 120K, and 1.2M hospitalization events) and tested the PHeP using a range of different GPUs (i.e., NVIDIA Tesla T4 16GB, NVIDIA P100 16GB, NVIDIA A100 40GB, and NVIDIA GraceHopper Superchip GH200 with H100 100GB).

Table 1 shows the performance in terms of execution time for pre-training and fine-tuning, indicating that the PHeP system successfully leveraged the available GPU resources, demonstrating faster pre-training and fine-tuning times with more advanced GPUs.

**Table 1:** BERT running time. The table shows the time spent, per epoch to, pre-train (PT) and fine-tune (FT) using different GPUs and input dataset sizes (the number of hospitalisation events N is reported).

|  | N=12K |  | N=120K |  | N=1.2M |  |
| --- | --- | --- | --- | --- | --- | --- |
|  | PT | FT | PT | FT | PT | FT |
| T4 | 15min 39sec | 13min 05sec | 2h 43min | 2h 20min | 28h 30min | 23h 45min |
| P100 | 9min 45sec | 8min 26sec | 1h 53min | 1h 24min | 19h 05min | 14h 00min |
| A100 | 4min 43sec | 3min 35sec | 45min 31sec | 40min 12sec | 7h 48min | 6h 03min |
| H100 | 1min 30sec | 1min 20sec | 15min 30sec | 14min 00sec | 2h 49min | 2h 10min |

The second experiment allowed us to test the *Prediction Environment*, a central component of PHeP. We pre-trained and stored two BERT models within the TrustAlert *Data Repository* to predict patients’ re-hospitalization at three months. The models were trained on a simulated cohort of 680 elderly patients (aged *>*65 years) with 12,161 unevenly spaced hospitalizations over 14 months. The dataset included a balanced number of female (N=6,039, 49.7%) and male (N=6,122, 50.3%) patients, with a median age of 78 years old (interquartile range (IQR): 73-83). The number of hospitalizations per patient ranged from 1 to 189 (median: 13, IQR: 8-20). Each hospitalization was accompanied by at most six diagnoses (median: 3, IQR: 1-4), encompassing 850 unique ICD-9 codes, with frequencies ranging from 1 to 1,260 (median: 18, IQR: 11-36). The most frequent diagnoses and recorded via ICD-9 codes were connected with cardio-metabolic (e.g., atrial fibrillation, benign essential hypertension, diabetes mellitus) and respiratory (e.g., acute respiratory failure, obstructive chronic bronchitis with (acute) exacerbation) diseases, in line with an ageing population. In 70% of the medical histories, the next hospitalization event happened within three months.

We observed an accuracy of 70,6% and 86.8%, for the *baseline* and the *English-HAD* models, respectively, probably due to the limited amount of data available to train the *baseline* model from a blank state, while the *English-HAD* benefitted from the pre-set weights.

These pre-trained models can be used throughout an intuitive and reactive web interface (Figure 1C, http://trustalert.hpc4ai.unito.it:3000/), where users should only upload a CSV file containing the patients’ hospitalization history. The results of the computation is a text file whose first column contains the patient ID, as provided in the input file, and the second column contains the prediction label, *i.e*., ‘1’ if the patient is expected to be re-hospitalised within three months, ‘0’ otherwise. In this way, the prediction tasks can be performed in minutes by non-experts without requiring any technical knowledge, software installation or parameter tuning, making predictive healthcare accessible to healthcare providers which often lack these capabilities. Notably, input files are not stored on our servers, thus respecting GDPR privacy regulations.

## 4 Conclusion

In this paper, we introduce the Predictive Healthcare Platform (PHeP), an innovative opensource platform developed under the TrustAlert project. PHeP is designed to simplify the use of pre-trained models in predictive healthcare, providing an intuitive web-based graphical interface. This makes it accessible to users without extensive computational skills or resources.

Specifically, in the reported case study, PHeP has demonstrated its efficiency and adaptability in handling large-scale hospitalization events. Our experiments, conducted with varying data sizes and different GPUs such as T4, P100, A100, and H100, have shown robust performance. The platform houses pre-trained BERT models that predict patients’ re-hospitalization at three months. These models, trained on a simulated cohort of elderly patients, achieved an accuracy of 70.6% and 86.8% respectively, validating the effectiveness of PHeP in predictive healthcare tasks. The platform’s user-friendly interface allows non-experts to perform these tasks in minutes, making predictive healthcare more accessible to healthcare providers. Looking ahead, future enhancements to PHeP will focus on expanding its range of capabilities, allowing, for instance, the prediction of the onset of new diseases and of the risk of non-urgent A&E accesses. We will also ensure that models are transparent and explainable, further improving its user-friendliness and accessibility. This positions PHeP as a valuable tool in the ongoing revolution of predictive healthcare.

## Data Availability

All data produced are available online at https://drive.google.com/file/d/1khP0nzleCArYQXxMYHmh7F-SIwiMC0G-/view

https://shorturl.at/xaecv

## Acknowledgments

D.B. is a Ph.D. student enrolled in the National Ph.D. in Artificial Intelligence, XXXVII cycle, health and life sciences course organized by Universita` Campus Bio-Medico di Roma.

## Availability of data and software code

The source code used to pre-train and fine-tune the model is available at: https://github.com/edoppiap/bert_medical_records. The source code of the service interface is available at: https://github.com/qBioTurin/TrustalertPredictionService. The simulated dataset used to train the BERT models is available at the following URL: https://shorturl.at/xaecv.

